# User Experience of a Rehab Community-Based, Affordable Robot Exercise System (Rehab CARES) for Gamified Motor and Cognitive Stroke Rehabilitation

**DOI:** 10.64898/2026.11.10.26367762

**Authors:** Reem M. Azar, Yiyang Shang, Erica L. Waters, Rochelle J. Mendonca, Alwyn Johnson, Paul Diefenbach, Pamela Z. Cacchione, Michelle J. Johnson

**Affiliations:** Rehabilitation Robotics Lab, University of Pennsylvania; Department of Bioengineering, University of Pennsylvania; Antoinette Westphal College of Media Arts and Design, Drexel University; RePlay Lab, Drexel University; Department of Mechanical Engineering & Applied Mechanics, University of Pennsylvania; Department of Rehabilitation and Regenerative Medicine, Columbia University; Recupero Robotics LLC; enAbleGames, LLC; School of Nursing, University of Pennsylvania; Penn Presbyterian Medical Center, Penn Medicine; Department of Physical Medicine & Rehabilitation, University of Pennsylvania

**Author notes:** **Corresponding author:** Reem Azar, Rehabilitation Robotics Lab, University of Pennsylvania, 1800 Lombard Street Philadelphia, PA 19146. Indicates equal contribution (Reem M. Azar, Yiyang Shang, Erica L. Waters).

**Keywords:** Rehabilitation Robotics, Stroke, Usability, Cognitive Rehabilitation, Motor Rehabilitation

## Abstract

**Purpose:** Robot-assisted therapy predominantly targets motor impairment, although post-stroke motor and cognitive impairments co-occur. We report user experience in a longitudinal deployment of a Rehab Community-based Affordable Robot Exercise System (RehabCARES) for delivering personalized motor/cognitive training.

**Methods:** Six adults with chronic stroke (FM-UE 20–61; MoCA 22–28) completed 12 therapy visits with THEAbot, a reconfigurable 1-DoF robot. Rehab CARES also included a haptic/biometric dynamic difficulty adjustment controller and eight games characterized by cognitive processes and motor/cognitive demand. We report gaming preference, usage, and user experience (pain, exertion, usability, affect, workload) across therapy.

**Results:** User experience varied, but generally, self-reported pain was low, perceived exertion was light to moderate, enjoyment was strong and stable, and sense of control increased while excitement plateaued. Workload showed decreasing frustration and effort. Game selection during therapy was partially consistent with motor/cognitive clinical profiles; preferences and perceptual abilities may have influenced choices. Usability scores exceeded acceptability at all time points, peaking at the final visit (86 ± 4.5 out of 100).

**Conclusions:** RehabCARES provided personalized motor/cognitive training to a heterogeneous cohort in a community-based setting, delivering a positive user experience. Hardware reconfigurability and motor/cognitive game variety likely enabled this success.

**Trial registration:** ClinicalTrials.gov, https://clinicaltrials.gov/study/NCT05542121, NCT05542121.

## Introduction

At one year post-stroke, 40% of stroke survivors experience moderate to severe disability ^1^. This disability can result from a combination of stroke-related motor and cognitive impairments. Specifically, the literature suggests that cognitive impairment occurs in about 60% of stroke patients, whereas motor impairment occurs in 61.7% of patients, with some reports indicating higher percentages of cognitive impairment (up to 80%) ^2–4^. Despite these poor outcomes, only 12% of people with stroke receive occupational therapy between 6 and 12 months post-stroke in the US ^5^. This need is even more pronounced in low and middle income countries (LMICs), which face access barriers including high costs, lack of trained clinicians, and difficulty accessing clinics ^6,7^. Community-based rehabilitation (CBR) can help to fill these gaps by bringing services to local communities and lowering the cost of care ^8,9^. Beyond increased accessibility, CBR has been shown to have benefits in physical and psychosocial domains ^10,11^. The sense of community instilled by CBR programs may also improve patient motivation, which correlates with improved rehabilitation outcomes ^12,13^.

Robot-assisted therapy presents an opportunity to improve post-stroke CBR and can provide comparable levels of post-stroke motor recovery as standard of care ^14^, while offering benefits such as increased therapeutic dose ^15^, increased motivation and engagement through gamification ^16,17^ and personalized rehabilitation ^18^. Beyond motor training, robot-assisted therapy may improve quality of life, cognition, and psychosocial function ^19^. Robot-assisted therapy can even be designed to support simultaneous motor and cognitive training ^20^. Despite the potential for rehabilitation robots to provide cognitive training, most robot-assisted therapy studies focus solely on motor training while cognitive training remains relatively underexplored.

Many robot therapy devices use the “assist-as-needed” control paradigm to adjust for multiple different levels of motor impairment ^21^, but this approach only accounts for motor impairment and may ignore adjustments for differences in cognitive function. Dynamic Difficulty Adjustment (DDA) addresses the need to adjust for cognitive levels by modulating each individual’s game parameters in real time, keeping each user near the “challenge point” that maximizes motor learning ^22^. Most rehabilitation DDA systems rely solely on game performance metrics to drive adaptation ^23^, but performance-only approaches can misattribute difficulty when sensory barriers, fatigue, or attentional lapses degrade scores without reflecting true motor capacity, a problem compounded when a therapist’s attention is distributed among multiple patients at once. Biocooperative control supplements performance with physiological signals such as heart rate and galvanic skin response ^24^, though integrating these in stroke populations is complicated by medication-altered baselines and signal noise.

The Rehab Community-based Affordable Robot Exercise System (Rehab CARES) is a robot gym concept that utilizes robotic stations focused on distinct motor and cognitive skills to enable a single therapist to oversee multiple patients in a collaborative therapy gym environment ^25^. The newest version of the Rehab CARES gym consists of three reconfigurable 1-DoF end-effector robot called THEAbot (Figure 1) ^26^ (Recupero Robotics LLC, Philadelphia PA), integrated with a gaming suite and novel hierarchical haptic/biometric DDA controller (enAblegames™) ^27^. We demonstrate the system’s potential for simultaneous motor and cognitive training by defining the cognitive process utilized in the games in our rehabilitation library and characterizing those games according to their motor and cognitive challenge. Together, these features are intended to support personalization to individual participant needs via reconfigurable hardware and a library of games with varying motor and cognitive demand. We present usability, acceptability, and utilization of a longitudinal, community-based deployment of Rehab CARES. We evaluate the system with user-reported surveys and individual utilization, including robot position and game selection over 12 therapy sessions, from a small, heterogeneous cohort of people with stroke.

**Figure 1.**
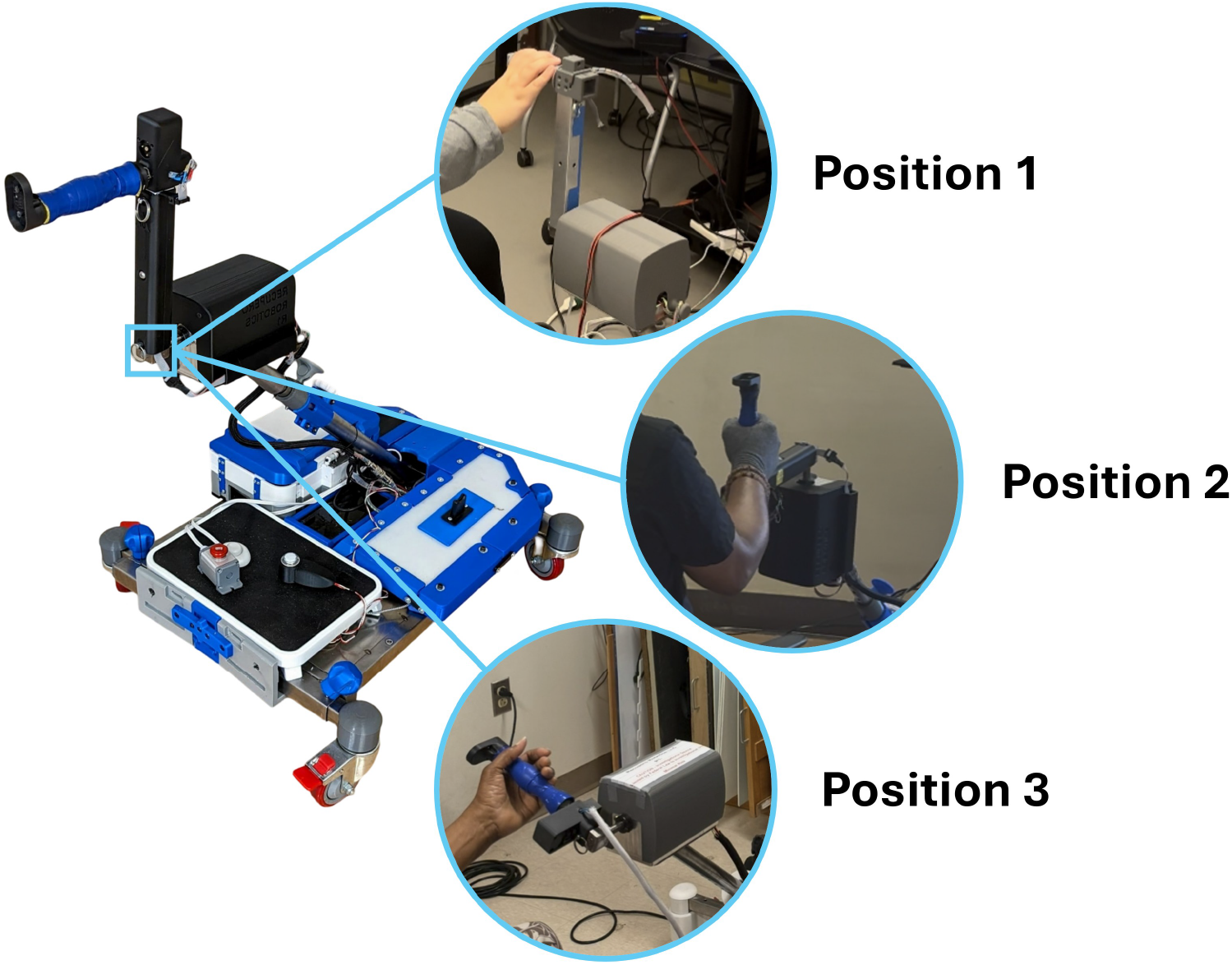
THEAbot is a 1-DoF reconfigurable robot with an interchangeable mounting hub at the drive shaft, which enables three configurations.

## Related Work

Conventional robot-assisted therapy pairs a single user with a single device under direct therapist supervision, but this configuration scales poorly and can increase costs, which is especially challenging in low-resource settings ^28^. When configured as a robot gym with multiple stations to accommodate group therapy for users with different therapeutic goals, rehabilitation robots can enable individualized practice while delivering CBR benefits such as reduced cost and sense of community ^25,28–30^. Several different robotic gyms for post-stroke arm rehabilitation have been presented in the past. Buschfort et al. showed that an “arm studio” consisting of six devices that trained different upper limb movements could increase intensity of inpatient subacute stroke rehabilitation ^31^. Aprile et al. furthered these results with a randomized control trial of inpatient subacute stroke patients, demonstrating that a gym consisting of four devices led to similar motor improvements as conventional therapy and increased the number of patients that a therapist could oversee ^29^. Bustamante-Valles et al. first demonstrated feasibility of robot gyms for chronic stroke rehabilitation in a low-resource setting ^30^. The robot/computer-based gym was also the first robot gym to explicitly focus on cognitive impairment by incorporating a “brain training” game station and inspired the evolution of the Rehab CARES platform as a more affordable robot gym ^25^. The current version of Rehab CARES builds on the robotic gym through three key features: reconfigurable hardware, gamified tasks, and adaptive difficulty adjustment. Together, these features help ensure that the system provides affordable, engaging, and personalized motor-cognitive training.

### Reconfigurable Hardware – Affordability and Comprehensive Motor Training

A key limitation to clinical deployment of rehabilitation robots is the high start-up cost ^18,32^. Decreasing the degrees-of-freedom (DoF) can dramatically reduce the cost of robot hardware ^33^. Low-DoF systems also do not lead to lower performance. In fact, end-effector robots offer equivalent or superior clinical outcomes for upper limb training. Lee et al. conducted the first direct randomized controlled trial comparing end-effector and exoskeleton architectures in chronic stroke patients, finding end-effector robots significantly outperformed exoskeletons on activity and participation outcomes ^34^. Likewise, meta-analysis of 31 RCTs similarly ranked end-effector robot-assisted therapy highest for improving upper limb motor function ^35^. Low-DoF end-effector robots, however, can limit the types of motion patterns a user can practice.

Reconfigurable robots may provide a solution, enabling multiple movement patterns while keeping hardware complexity and costs low. Examples of 1-DoF reconfigurable rehabilitation robots include the Bi-Manu-Track (bimanual forearm pronation/supination and wrist flexion/extension) ^36^ and the CR2-Haptic (four different grasps; forearm pronation/supination, wrist-flexion/extension, wrist ulna-radial deviation, and elbow and shoulder flexion/extension) ^37^. These standalone devices demonstrate that reconfigurable hardware can enable a single device to train multiple movements. In our previous work, we validated that our 1-DoF, reconfigurable robot, THEAbot, can provide three different types of movement (elbow and shoulder flexion/extension, shoulder abduction/adduction, and forearm pronation/supination) ^26^. These versatile robotic systems enable comprehensive arm exercise on a single platform.

Unlike previous robot gym iterations, Rehab CARES uses a single device, THEAbot, for all stations. Because the robot enables three different arm exercises, clinics do not need to purchase separate systems for each type of movement. This reconfigurable hardware reduces start-up costs by enabling clinics to slowly grow the number of gym stations without compromising on therapeutic versatility. Furthermore, a robot gym with a single robotic platform can alleviate the administrative burden of training clinicians on new equipment. Rather than learning to operate three different devices, clinicians can train on a single system. Finally, a reconfigurable system is more robust to changing therapy needs, allowing users to practice whichever movement best meets their goals, regardless of how the other gym stations are utilized.

### Gamification – Engagement and Cognitive Training

Gamification can support engagement in rehabilitation by restructuring repetitive movement practice as goal-directed activity with explicit objectives and immediate feedback. In post-stroke robot-assisted therapy, a meta-analysis of 42 randomized controlled trials involving 1,760 participants found that serious-game interventions produced greater improvements than conventional therapy in upper-limb function, activity and participation, with the largest pooled effect observed for participation ^38^. Pixa et al. identify an appropriate balance between challenge and skill, clear goals, and immediate feedback as design characteristics associated with engagement in post-stroke rehabilitation games ^39^.

A separate body of work considers rehabilitation games not only as motivational structures for motor practice but also as vehicles for *cognitive* practice.

This distinction is particularly relevant after stroke, where motor and cognitive impairments frequently cooccur and cognitive status can influence responsiveness to motor rehabilitation ^40^. Gameplay that requires rapid color discrimination, maintenance of a target sequence in working memory, divided attention, or strategic evaluation of competing goals recruits cognitive processes in addition to those involved in producing the required movements. Consequently, serious games have been investigated as interventions for attention and related cognitive functions. Reviews report improvements in at least one outcome related to attention in a majority of the studies examined, while also identifying persistent methodological limitations, including small samples and heterogeneous outcome measures ^41^. Two games that elicit similar joint movements may therefore differ substantially in their attentional, mnemonic, perceptual, or executive demands.

Despite this interaction between motor and cognitive demands, rehabilitation games are commonly described primarily in terms of their physical requirements, such as the body segments involved, movement range, repetition, or resistance. When cognitive characteristics are reported, games are often categorized by genre or by the broad cognitive domain they are intended to address rather than by an explicit, graded characterization of the cognitive demands imposed during gameplay ^41^. Similarly, difficulty is generally represented as progression within an individual game rather than as a common dimension that can be compared across games in a library.

This creates an asymmetry in game selection. A clinician can often assess whether a game is compatible with a particular motor goal from the game’s physical requirements, whereas selecting a game for a particular cognitive profile may depend more heavily on prior experience, informal judgment, or trial and error. This limitation is particularly relevant in rehabilitation settings in which individuals with heterogeneous motor and cognitive abilities train concurrently and therapist attention is distributed among multiple patients or stations ^25,29^. In such settings, a clinician may need to alter cognitive demand while preserving a desired movement pattern—for example, by selecting an activity that elicits a comparable physical action while placing lower demands on attention, working memory, or executive control. Descriptions centered primarily on physical requirements provide limited support for this form of selection.

### Dynamic Difficulty Adjustment – Personalization and Progressive Challenge

Individualized difficulty in rehabilitation derives from two complementary frameworks: Csikszentmihalyi’s Flow Theory, which identifies challenge-skill balance as the determinant of optimal engagement ^42^, and the Challenge Point Framework (CPF), which holds that the optimal challenge point must be continuously updated as skill develops ^22^. Pixa et al. confirmed that challenge-skill balance, clear goals, and immediate feedback are the conditions most reliably associated with sustained engagement in stroke rehabilitation ^39^.

Early DDA implementations relied on performance metrics alone; while these effectively capture aggregate task outcomes (such as trial success or completion time), they provide delayed feedback that only registers after a movement concludes, failing to capture the real-time divergence between apparent success and underlying physiological distress ^43^. Novak et al. showed that fusing biometrics with performance increased the accuracy of the controller from approximately 60% to 82% ^24^. Our Partially Ordered Set Master (POSM) algorithm operationalizes the CPF in real time, computing individual challenge points from physiological and kinematic streams at each station and continuously adjusting both virtual game parameters and physical robot resistance.

Biocooperative control, in which the robot’s control loop is closed on the user’s physiological and psychological state (e.g., heart rate, galvanic skin response, affect) rather than purely on position or force, has emerged as a strategy for adapting rehabilitation intensity to their internal state in real time ^44^. Rodriguez-Guerrero et al. presented the closest single-station precursor to our gym architecture, simultaneously adapting game difficulty and haptic resistance from HR and GSR signals within a Flow-theoretic framework ^45^. Our architecture extends this to a multi-station gym by treating biometrics as a safety constraint, triggering protective difficulty reductions when physiological signals conflict with performance data, rather than as a continuous ground truth signal. Gandolla et al. validated a parallel approach at the hardware layer, demonstrating that closing the control loop based on active participation metrics significantly increases physical workload and affect ^46^.

## Methods

### System Design

*Robot* THEAbot is a 1-DoF, reconfigurable robot designed for upper limb rehabilitation (Figure 1). The robot consists of a quasi-direct drive (QDD) actuator, which combines a high torque density motor, gearbox, encoder, and motor driver ^47^, connected through the drive shaft to a mechanical torque limiter that limits torques to 30*Nm*. An interchangeable hub attached to the drive shaft allows for easily interchangeable handles. The robot is mounted on a height-adjustable mobile frame that accommodates standard chairs and wheelchairs. The robot can be reconfigured into three distinct positions that elicit different movements and muscle activation patterns: Position 1 (shoulder and elbow flexion/extension), Position 2 (shoulder abduction/adduction), and Position 3 (forearm pronation/supination) ^26^.

The system is operated by microcontroller, which controls the actuators and connects to the force sensing robot handles and the bluetooth enabled “biopack,” which is strapped to the user’s arm and monitors heart rate, galvanic skin response, and trunk movement via inertial measurement units. The controller resists the user’s movement by applying force opposite to the user’s velocity. The gains of the controller were selected to apply the appropriate range of forces based on the three positions of the robots at eight different difficulty levels.

### Software Architecture

#### Unified Kinematic Input

Games interface with THEAbot through SUKI+, a sensor-agnostic middleware framework that connects physical input devices (e.g., full-body tracking, camera pose-tracking, and serial robotic devices) to game applications without requiring game-code modifications ^48^. SUKI+ abstracts motion data via a *schema* that maps physical movements to standard game primitives (signals, Booleans, scalars, and 2D/3D vectors) and an *extents* mechanism that normalizes each user’s usable range of motion to a [0, 1] interval. This allows THEAbot’s single-degree-of-freedom rotary position to drive diverse game controls, with its three physical configurations exposed as distinct tracking modes. Input extents can be personalized per session: initial values combine observed input with historical portal data weighted by a therapist-configured default percentage, and subsequently adapt in real time to the user’s active movement range.

#### Dynamic Difficulty Adjustment

Game difficulty is governed by the Haptic/Biometric-Based Dynamic Difficulty Adjustment (HBB-DDA) framework, which uses a modified Partially Ordered Set Master (POSM) algorithm to update a belief distribution over difficulty states based on performance evidence ^27,49^. Extending prior single-station validation ^27^ to a multi-station environment, HBB-DDA adapts at each step using game-specific performance signals alongside heart rate and galvanic skin response deviations from a same-day resting baseline. Each game defines distinct performance signals and discretized difficulty dimensions that adapt independently (Table 1), supplemented by THEAbot’s viscous resistance as a shared physical difficulty dimension. Prior to each session, therapists configured robot orientation, initial difficulty, default extent percentage, duration, and (for Sky-Burger) ingredient-fall speed. An available robotic assistive mode for four games was disabled throughout this study.

**Table 1.**
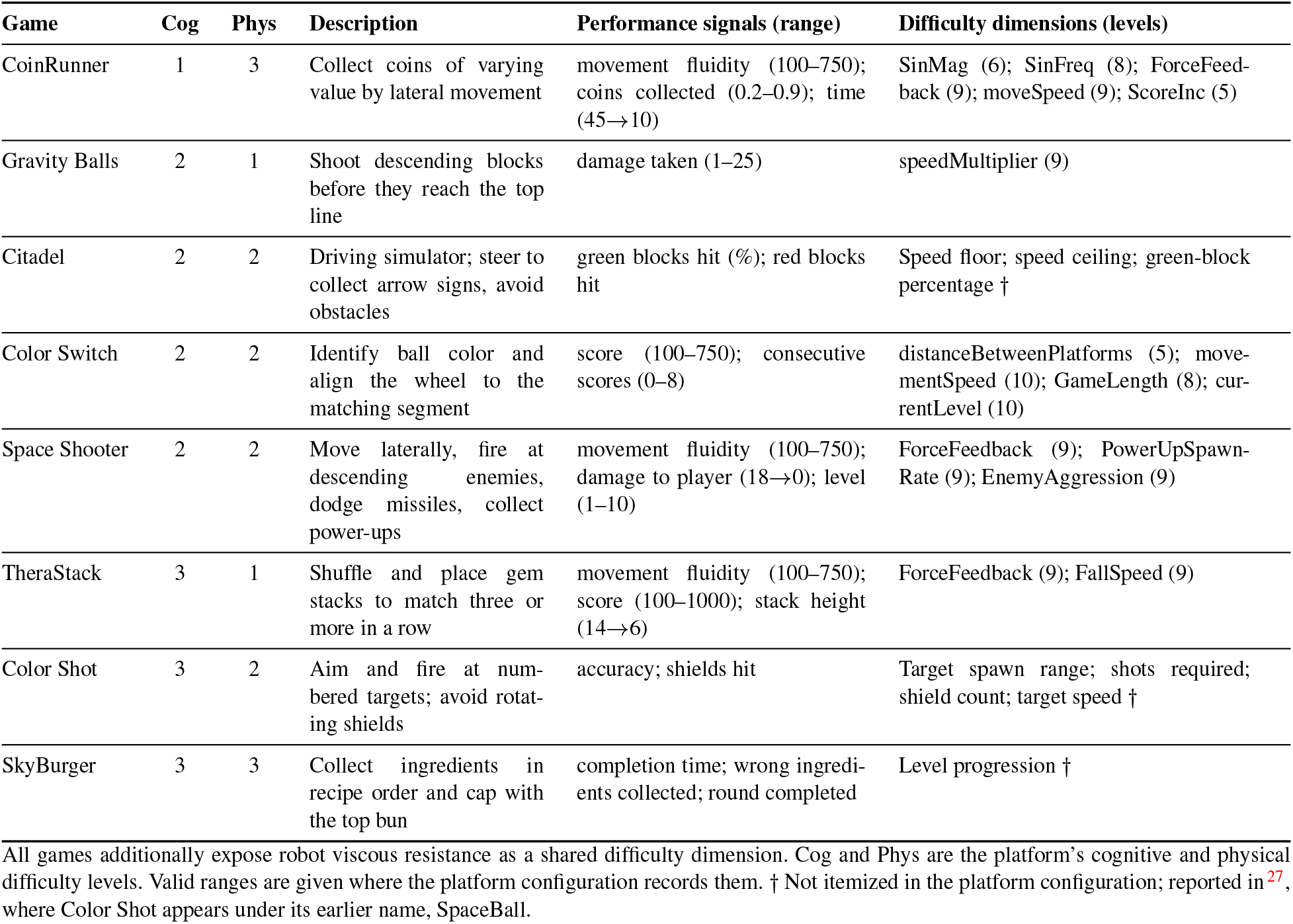
Therapy games, difficulty levels, and dynamic difficulty adjustment configuration.

### Therapy Games

Eight single-player enAblegames™ therapy games were deployed via SUKI+ and HBB-DDA, spanning varied motor and cognitive demands (Table 1).

#### Cognitive Processes

All games were retrospectively characterized as a repeating sequence of two (CoinRunner) to five (TheraStack) cognitive processes across seven categories (Figure 2): *Spatial* (locating objects relative to the player); *Spatial-Motor*, split into *SM-1* (vertical/horizontal alignment) and *SM-2* (precise angular alignment); *Dexterity-Motor* (discrete timed actuation during continuous movement); *Strategic (1)* (binary pursue/avoid decisions); *Strategic (2)* (rule-conditional decisions, e.g., recipe order); and *Spatial-Strategic* (multi-variable spatial analysis).

**Figure 2.**
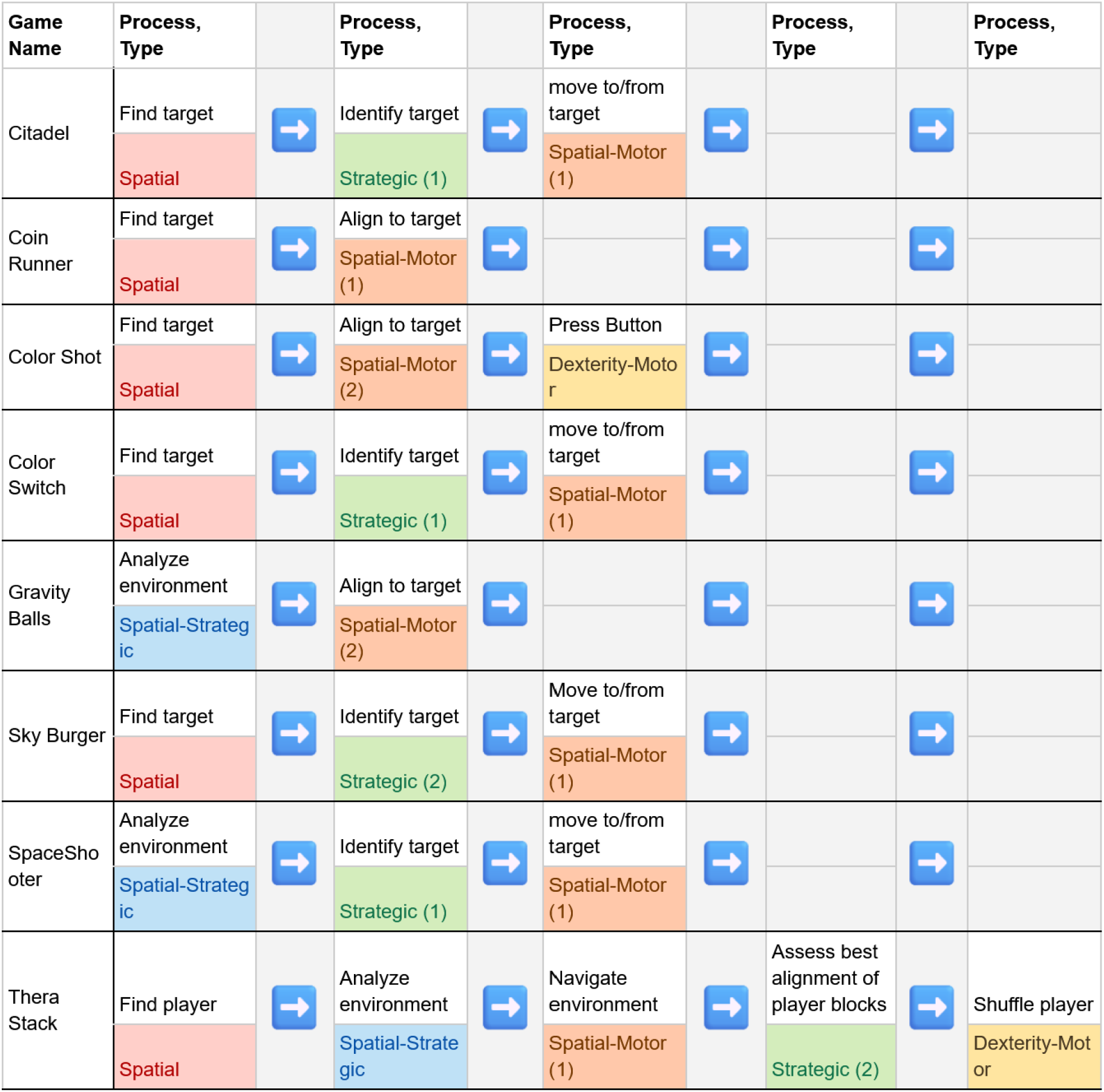
Therapy games broken down into cognitive sequences, ranging from simple chains like CoinRunner (Spatial →SM-1) to five-stage sequences in TheraStack.

#### Cognitive and Physical Difficulty Levels

Games are rated on cognitive (Cog, 1–3) and physical (Phys, 1–3) difficulty levels, which serve as clinician selection filters in the game launcher. A game’s Cog rating reflects the highest process tier in its sequence rather than sequence length: Tier 1 (*Spatial, SM-1*), Tier 2 (*Strategic (1), Spatial-Strategic, SM-2*), or Tier 3 (*Strategic (2), Dexterity-Motor*). Phys ratings reflect required gross and fine motor demands. Across the 3 *×* 3 difficulty grid, the eight games occupy six cells (Figure 3). Because the axes operate nearly independently (e.g., TheraStack is Cog-3/Phys-1; CoinRunner is Cog-1/Phys-3), clinicians can vary cognitive and physical demands orthogonally, though the library currently lacks Cog-1/Phys-2 coverage.

**Figure 3.**
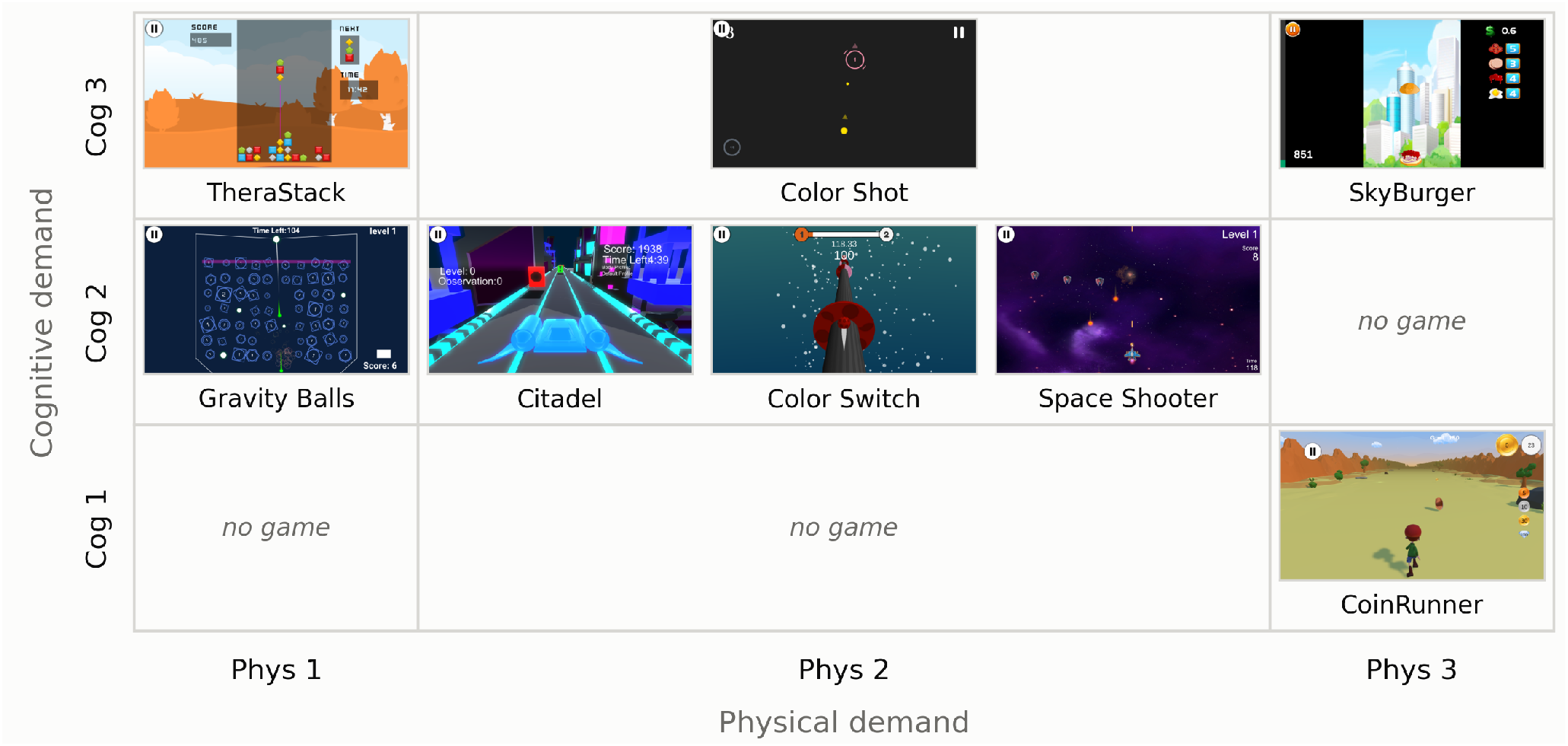
Coverage of the therapy game library across cognitive and physical difficulty levels (occupying six of nine cells).

### User Study

#### Clinical Environment

Data were acquired at two separate clinical sites. The first site was a community-based site called Mercy LIFE (Living Independently for Elders) West Philadelphia, which is a Program of All-Inclusive Care for the Elderly (PACE) that offers comprehensive medical and social services to nursing home–eligible older adults living in the community. PACE participants are generally dually eligible for Medicare and Medicaid. The second site was at the Rehabilitation Robotics Lab (RRL) of University of Pennsylvania. All therapy visits were overseen by an occupational therapist (OT), and all therapeutic decisions were made by an OT. Vitals such as heart rate, blood pressure, and oxygen levels were collected at the beginning and end of the visit.

#### Participants

All participants were at least 55 years old with a documented diagnosis of stroke that occurred at least 3 months prior to study enrollment. Hemiplegia was confirmed using the Fugl-Meyer Upper Extremity Assessment (FM-UE), with scores *<*62 indicating motor impairment. Participants with mild to moderate levels of cognitive impairment were included. Participants were required to have completed either a CT or MRI scan after stroke onset. Written informed consent was obtained from all participants prior to study participation.

#### Clinical Assessment

After being recruited, participants were consented and assessed using standard clinical and robot-based assessments. Participants completed several motor and cognitive assessments, including the Fugl-Meyer Assessment (FM-UE) upper extremity ^50^ and the Montreal Cognitive Assessment (MoCA) ^51^. Participants also completed three robot-based assessment games to evaluate their motor and cognitive abilities. More information on the complete clinical battery is reported in the clinical trial listing for the Rehab CARES study (phase II) ^52^.

#### Pre-Therapy Game Evaluation

The first robotic therapy game visit was led by an OT so they could understand the participant’s functional abilities and limitations, and ensure that they were safe to use the robot with minimal supervision for future robotic sessions. To best gauge functional range per game, participants played each game in robot Position 2 for three minutes. Within this time, the OT observed how the participant performed and interacted with both the robot and the game. At the end of each game, participants were asked about their enjoyment (“Did you like playing this game?”) and interest (“Are you interested in playing this game again?”). At the end of the first visit, participants were also asked to rank the games according to preference, selecting the games using four superlatives (“First Favorite”, “Second Favorite”, “Third Favorite”, “Least Favorite”).

#### Therapy Visits

Prior to beginning any visit, the participant’s blood pressure, pulse oxygen level, and heart rate were measured. If the participant’s vitals were within normal limits (as defined by their on-site nurse), the participant continued to the therapy visit. Each participant was equipped with the biopack and seated at the robot. Individual game sessions lasted 15–30 min, with two to five sessions completed during a 60-min therapy visit. After each session, participants were asked to rate their pain on a 10-point visual analog scale (VAS) and their exertion on a 10-point Borg scale. The OT selected the starting game and position using a client-centered approach combining the participant’s preference with their own recommendation informed by observed physical and cognitive abilities, therapeutic goals, and day-to-day factors such as fatigue and pain 1.

#### Usability Surveys

The OT conducted three surveys to assess the user’s experience with the device at the first, sixth, and twelfth visits (beginning, half-way, and at the end of the therapy visits). The *NASA-Task Load Index* (NASA-TLX) assesses the user’s workload in seven domains: mental demand, physical demand, temporal demand, performance, effort, and frustration ^53^. A lower score indicates a lesser workload/frustration or better performance. Lower scores on performance and frustration domains are desirable, while moderate effort and workload would indicate sufficient challenge in the therapy tasks. The second survey conducted was the *System Usability Scale* (SUS), which is a 10-item questionnaire focused on measuring perceived usability of the robotic system. SUS is scored out of 100 points with a score of 68 or higher indicating acceptable usability ^54^. Lastly, the *Self-Assessment Manikin* (SAM) is a non-verbal questionnaire that asks the user’s emotional response to the system on a 9-point scale, specifically in engagement, excitement, and confidence domains ^55^.

### Data Analysis

Given the small sample size and focus on usability and user experience, we focus on descriptive statistical analysis. Specifically, we report total responses to the pre-therapy game evaluation. Likewise, as game and robot position exposure was neither randomized nor balanced, differences in game use may reflect participant characteristics, therapist judgment, participant preference, scheduling, or other unmeasured factors. We therefore make no inferential comparisons between games and instead report descriptive statistics across participants per game and position. For DDA analysis, we describe sampled sessions from a single participant; the full body of therapy logs remains unprocessed, so these trajectories illustrate the controller’s per-game behavior rather than characterizing it across the cohort. Finally, we report descriptive statistics for each usability survey across the three survey time points. These survey results are interpreted according to established scoring definitions and relative changes across sessions.

## Results

A total of 6 participants (Age: 69.3 *±* 7.4 years; 50% female, 50% male; 100% Black) completed the game therapy using the THEAbot. Table 2 summarizes the demographics and clinical characteristics of the participants. One participant (024) had mild motor impairment (FM-UE *>* 50) and no cognitive impairment (MoCA ≥26). Four participants (006, 018, 022, and 028) had mild motor impairment and mild cognitive impairment (18 ≤ MoCA *<* 26). One participant (029) had severe motor impairment (FM-UE ≤22) and mild cognitive impairment. At Mercy LIFE, four participants (006, 018, 022, and 024) averaged 2 therapy visit per week for 6 weeks while at Penn Medicine Rittenhouse, the two participants (028 and 029) averaged 3 therapy visits per week for 4 weeks. Participant 029 only completed 11 visits due to his physical limitations from stroke (FM-UE ≤ 22).

**Table 2.** Participant Demographics and Clinical Characteristics.

| **ID** | **Age Range (yrs)** | **Sex (M/F)** | **Education (yrs)** | **Time Since Stroke (yrs)** | **FM-UE (/66)** | **MoCA (/30)** |
|---|---|---|---|---|---|---|
| 006 | 60-65 | M | ≤ 12 | 3 | 55 | 23 |
| 018 | 65-70 | F | ≤ 12 | 3 | 58 | 25 |
| 022 | 65-70 | M | > 12 | 1 | 52 | 22 |
| 024 | 75-80 | F | ≤ 12 | 3 | 61 | 28 |
| 028 | 75-80 | F | > 12 | 1 | 60 | 24 |
| 029 | 55-60 | M | > 12 | 1 | 20 | 24 |

### Pre-Therapy Game Evaluation

Overall, participants responded positively to most games. For Citadel, Color Shot, Gravity Balls, and Space Shooter, all participants responded “yes” to liking the game and yes to interest in playing the game again. All participants selected “yes” to liking Coinrunner, but one participant indicated that they were not interested in playing it again. One participant responded “no” to both questions for SkyBurger and Color Switch with all other participants selecting “yes.” Finally, two participants responded “no” when asked if they liked playing TheraStack, and three participants indicated that they would not want to play it again.

Figure 4 shows each participant’s first, second, third, and least favorite games. Game preferences varied, but notable trend include the overall positive response to Space Shooter, Color Switch, and Citadel and the mostly negative response to TheraStack. Gravity Balls, Color Shot, and CoinRunner received more mixed responses. Interestingly, Sky Burger was not selected for any of the superlatives.

**Figure 4.**
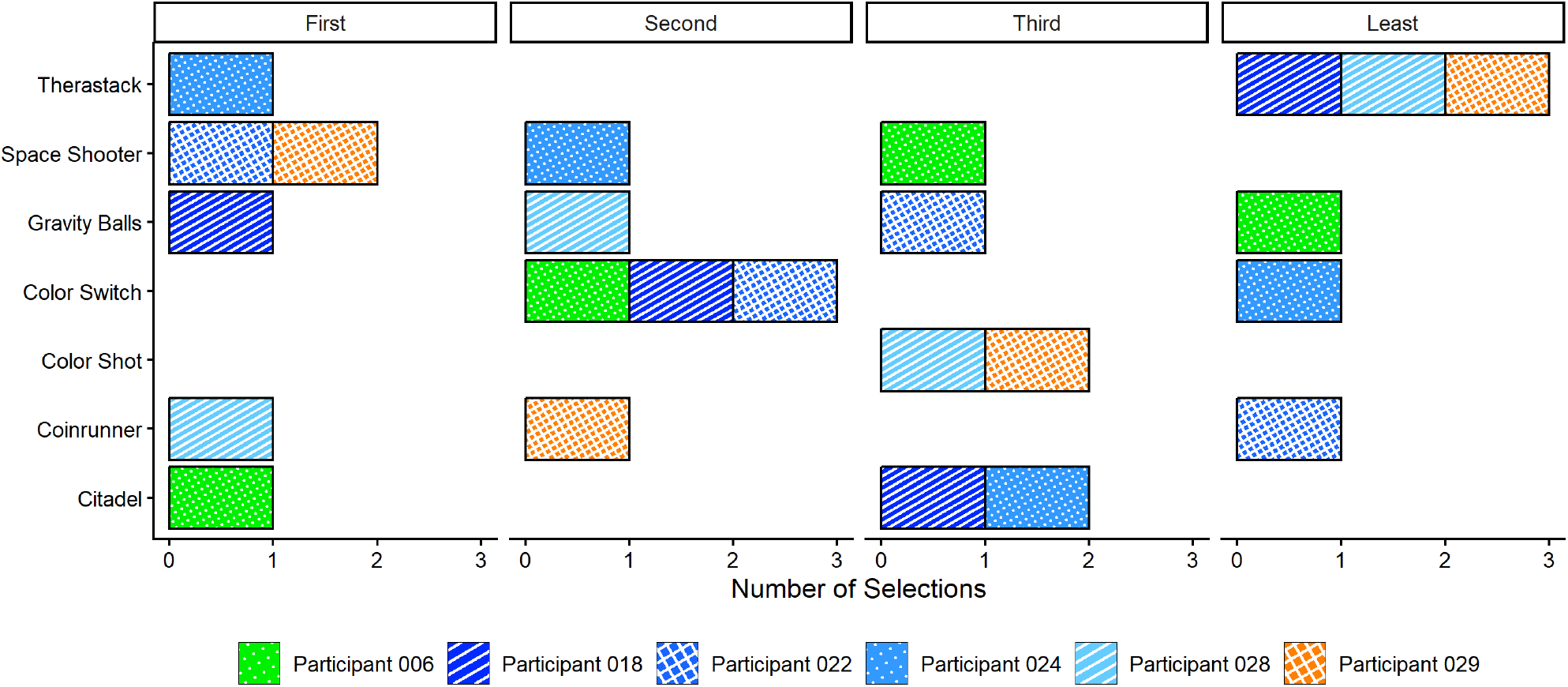
Game preference selections for first, second, third, and least favorite game. Note that colors correspond to impairment groups: green for mild motor impairment with no cognitive impairment, blue for mild motor and cognitive impairment, and orange for severe motor impairment with mild cognitive impairment.

### User Experience Across Therapy Visits Therapeutic Personalization of the Robotic Gym

This study’s participants represent a diverse range of clinical motor and cognitive profiles (Table 2). Similar to traditional therapy, we note that OT decisions in robot-assisted therapy was dynamic: adjusting to the participant’s overall goals, functional limits, preferences, and current status, e.g. fatigue and pain levels. For example, participant 029 had severe impairments in their upper extremity, specifically in their wrist and fingers. With the support of a glove, this user’s therapy focused on movements that involved more the wrist, e.g., Positions 2 and 3 (Figure 5B). For participants with mild to moderate motor impairments, however, the OT selected Position 3 less because it localizes more on wrist movements (Figure 5B). The OT also prioritized safety. Notably, participant 028 played mostly in Positions 2 and 3 not due to functional limitations in movement, but rather, their strength in Position 1 could shift the robot base during fast movements, presenting a potential safety issue. To minimize risk to both the participant and the device, the OT selected Positions 2 and 3 more frequently (Figure 5B).

**Figure 5.**
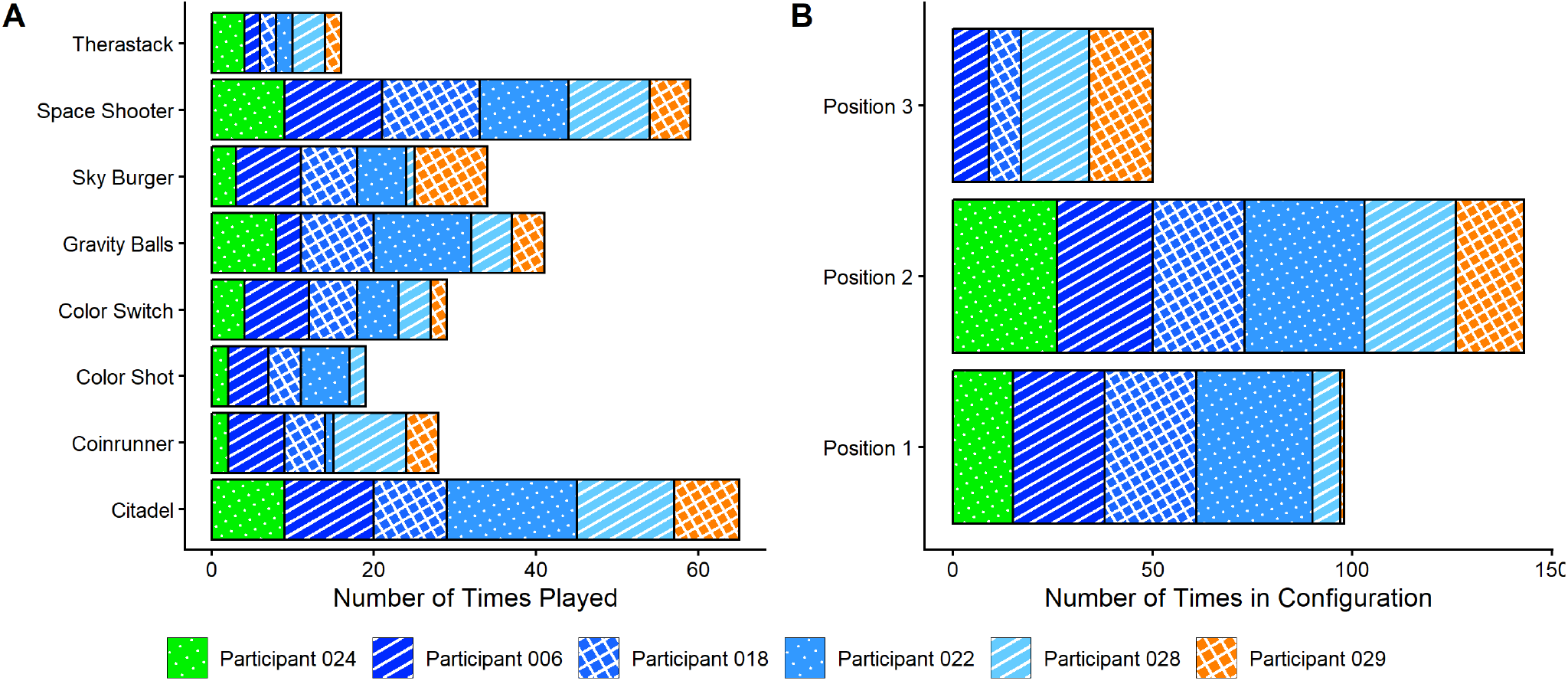
Therapy activity selections for game and robot position. Note that colors correspond to impairment groups: green for mild motor impairment with no cognitive impairment, blue for mild motor and cognitive impairment, and orange for severe motor impairment with mild cognitive impairment. **A:** Number of times each game was played across therapy. **B:** Number of times each position of the robot was experienced across therapy.

#### Pain

Self-reported pain was low with a mean VAS score of 0.30 *±* 0.92. Of the 256 sessions in which pain was reported, there were 24 instances of mild pain (0 *<* VAS *<* 4), 4 instances of moderate pain (VAS = 4), and 2 instances of greater than moderate pain (VAS *>* 4).

#### Exertion

Mean Borg perceived exertion across all sessions was 2.61 *±* 1.60. Per-game mean exertion was similarly light-to-moderate for Citadel (2.33 *±* 1.54), Gravity Balls (2.34 *±* 1.44), Color Shot (2.35 *±* 1.69), CoinRunner (2.50 *±* 1.29), Space Shooter (2.54 *±* 1.37), and Color Switch (2.77 *±* 1.54). Two games, Sky Burger (3.30 *±* 2.10) and TheraStack (4.14 *±* 2.32), produced moderate exertion.

#### Games Played

Figure 5A shows the number of times each game was played. Of the more widely used games (mean number of times played ≥5), Citadel (*Total* = 65, *mean* = 10.8 *±* 2.93) was the most popular game, followed by Space Shooter (*Total* = 59, *mean* = 9.83 *±*2.64). Of the less widely used games (mean number of times played *<* 5), TheraStack (*Total* = 16, *mean* = 2.67 *±*1.03) was the least popular game.

#### Robot Position

Figure 5B shows the number of times the robot was used in each position. Position 2 was the most common (*Total* = 143, *mean* = 23.8 *±* 4.26), followed by Position 1 (*Total* = 98, *mean* = 16.3 *±* 10.7) and Position 3 (*T otal* = 50, *mean* = 12.5 *±* 4.65).

#### DDA Adaptation

The DDA adapted independently across games for the same participant. In participant 006, Space Shooter level progressed from 10 →14→ 18 across three sampled sessions, while Citadel held steady at level 8, confirming that POSM operated on per-game performance signals rather than applying a global difficulty offset. Between-visit haptic resistance trajectories showed the same game-specific behavior: in Citadel, resistance ramped rapidly to near-ceiling levels within 100–150 seconds and was sustained, with ramp time decreasing across days 2, 7, and 11; in Space Shooter, resistance started at moderate levels and declined progressively within each visit. All of these are achieved while the scores remain stable, demonstrating that the DDA provides sufficient challenge. The two games produced qualitatively opposite resistance trajectories for the same participant on the same day. These two games produced opposite resistance trajectories for the same participant on the same day, which is the behavior the per-game difficulty configuration in Table 1 is designed to produce.

### Overall Usability Evaluation

#### System Usability Scale

Mean SUS scores were above the minimum acceptable score (68 out of 100) at both the first (77.5 *±*16.7) and second (70 *±* 18.0) survey points. THEAbot SUS scores were highest at the third and final survey point (86 *±* 4.54).

#### Task Load

Figure 6 shows NASA-TLX results for six domains of task load. Mean frustration was low to moderate at the first and second (24.2 *±*27.6, 36.7 *±*43.4) survey time points and dropped to very low at the third survey time point (10 *±* 14.6). Mean perceived effort was moderate at first (44.2 *±*27.5), with a slight decrease over time at the second and third survey time points (30.0 *±* 25.3, 27 *±* 24.1). Mean perceived performance, where a lower score indicates better performance, was low to moderate at all three survey time points (20.8 *±* 16.6, 30.0 *±* 20.5, 19.0 *±* 16.4).

**Figure 6.**
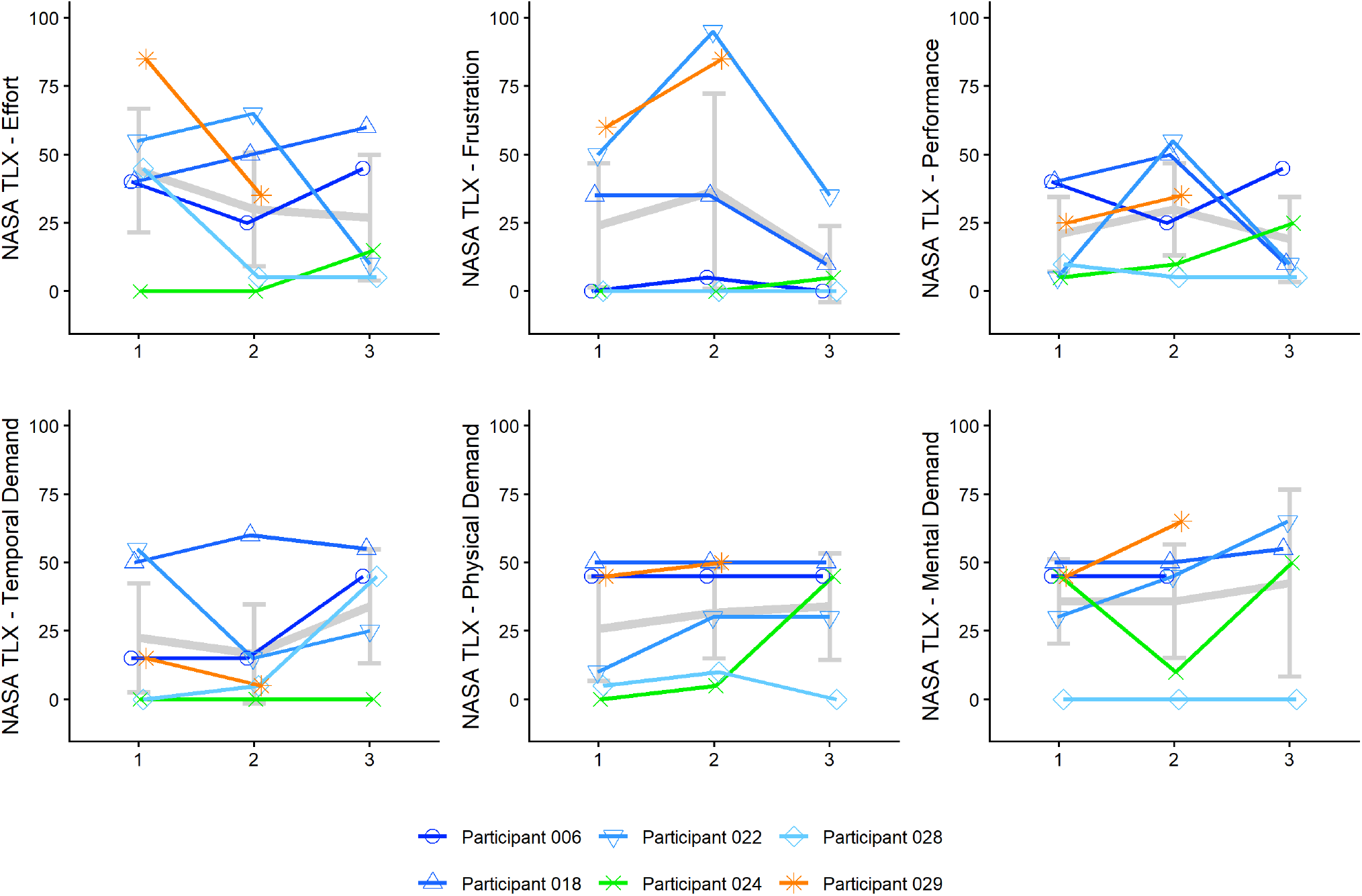
NASA Task Load Index in six domains at three survey points. Note that colors correspond to impairment groups: green for mild motor impairment with no cognitive impairment, blue for mild motor and cognitive impairment, and orange for severe motor impairment with mild cognitive impairment.

Mean temporal task load was similarly perceived as low to moderate at all three survey time points (22.5 *±* 24.2, 16.7 *±* 22.1), 34.0 *±* 21.9). Mean physical task load was perceived as moderate at first (25.8 *±* 23.1), with a slight increase at the second and third survey time points (31.7 *±* 20.2, 34.0 *±* 20.4). Finally, mean mental task load was perceived as moderate at all three survey time points (35.8 *±* 18.8, 35.8 *±* 25.2, 42.5 *±* 29.0).

#### Self-Assessment Manikin

Figure 7 shows SAM results for three emotional domains. Mean valence was generally positive or “happy” at all three time points (2.67*±*1.51, 3.0 *±*1.79, 2.6*±* 2.19). Mean arousal indicated that participants were initially “excited” by the system at the first survey time point (2.67 *±* 1.51), but their arousal shifted to moderately simulated for the later surveys (5.0 *±* 3.35, 5.0 *±* 3.16). The participants’ sense of control was generally high and increased with increased exposure (6.33 *±* 1.63, 7.33 *±* 2.66, 8.0 *±* 1.73).

**Figure 7.**
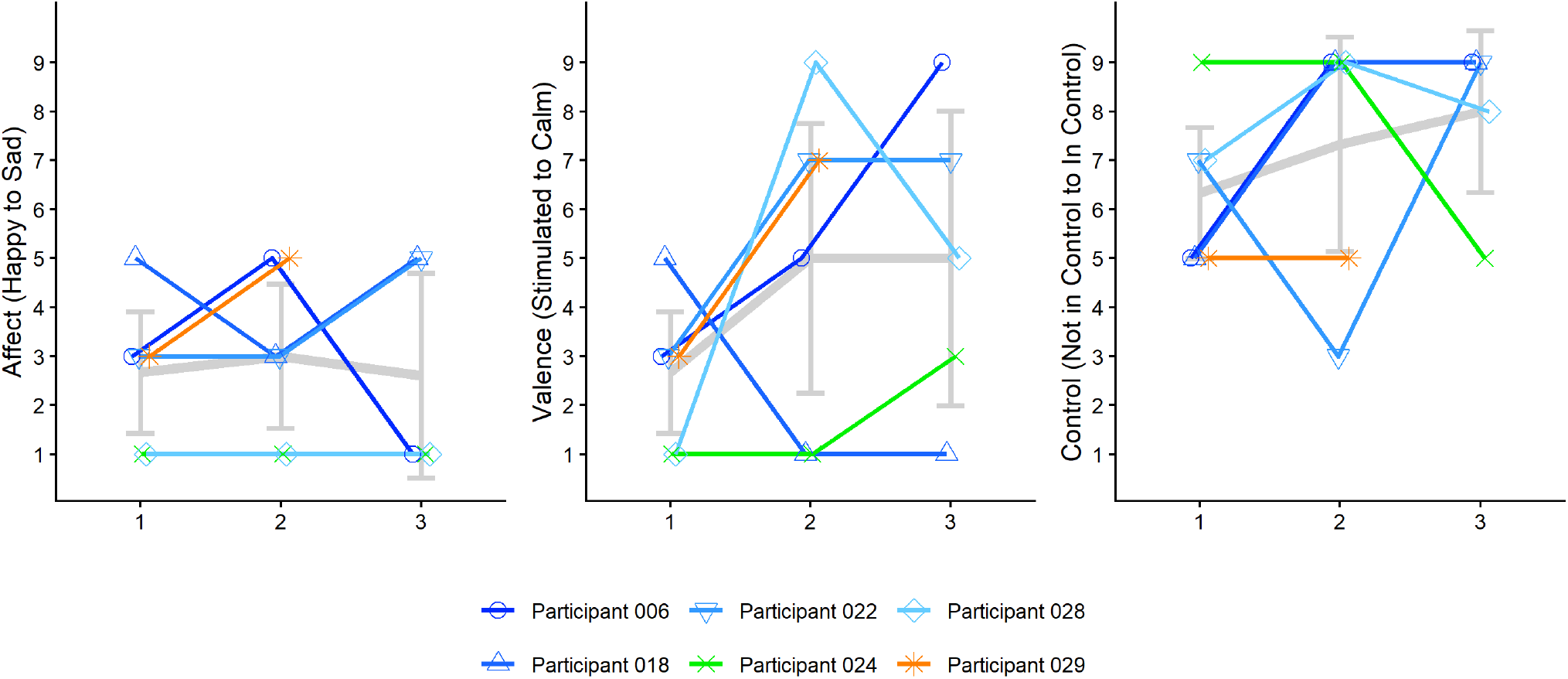
Self-Assessment Manikin in three domains at three survey points. Note that colors correspond to impairment groups: green for mild motor impairment with no cognitive impairment, blue for mild motor and cognitive impairment, and orange for severe motor impairment with mild cognitive impairment.

## Discussion

User experiences varied throughout the course of therapy, but in general self-reported pain was low with light to moderate exertion during game play with the robots. Game selection during therapy was partially consistent with clinically appropriate motor and cognitive demand. The system was overall acceptable with mean SUS scores above the acceptable cut-off score (68 out of 100). We observed increasing acceptance with increasing exposure to the therapy system. Engagement was consistently positive. Interestingly, we observed a potential novelty effect with SAM-arousal shifting from “excited” to moderately stimulated by the end of the therapy experience. Despite decreasing excitement as novelty wears off, the user experience appears to improve with time, as evidenced by increased usability, increased feelings of control, decreased perceived effort, and decreased frustration. Together, these results indicate that while initial novelty may wear off, the robotic gaming system is capable of sustaining usability and engagement over time.

### Selection of Games and Robot Configuration

The motor and cognitive abilities of the patients informed the selection of the game challenge, and selection frequency was broadly consistent with the cognitive characterization of each game. The four Cog-2 games (Gravity Balls, Citadel, Color Switch, Space Shooter) were selected more often on average than the three Cog-3 games (ThereStack, Color Shot, SkyBurger) (Figure 5A), while the single Cog-1 game (CoinRunner) was selected less frequently (Figure 5A). The fact that intermediate-demand games were selected the most often is consistent with the idea that mild cognitively impaired users desired games that challenged them at an intermediate level.

The characterization of games by their cognitive processes did not fully explain why and how often they were selected. For example, Citadel and Color Switch share the same cognitive sequence (Figure 2) and Cog-2/Phys-2 ratings, yet Citadel was selected much more frequently (Figure 5A) suggesting that participants’ preferences also plays a role (Figure 4). In other words, games may engage the same cognitive process and still not be played equally; the game design matters, depending on the participant’s personal preference. Therapist notes corroborated these findings with Citadel and Space Shooter noted as the most enjoyable and also the most frequently selected, while TheraStack was noted as the least enjoyable and was also selected the least. Perceptual accessibility may have also contributed; the therapist notes indicate that participants had difficulty discriminating task-relevant colors in CoinRunner, TheraStack, and Color Switch, and all three were less frequently selected.

Pre-therapy game preferences did not clearly associate with selection frequency and did not always remain static. For example, every participant reported enjoying Color Shot and wanting to play it again, yet, it was one of the least selected games (Figure 5A). Conversely, games that were the most highly rated during the pre-therapy evaluation, namely Citadel, Gravity Balls, and Space Shooter, were the most frequently played games in the library. In another example, participant 028 initially ranked TheraStack as her least favorite game (Figure 4). The therapist later observed that she came to enjoy it after additional play time and cueing, and she ended up playing TheraStack more often than the other participants (Figure 5A), indicating that preference changed with exposure.

Games with dual task demands were played infrequently. Color Shot and TheraStack are the only games whose cognitive sequence includes the dexterity-motor process, requiring timed actuation coordinated with movement of the same limb, and they were the two least frequently selected. Within the three Cog-3 games, the two games that required bilateral hand engagement with discrete push-button input from a participant’s less impaired limb were selected less often than the one that did not.

When selecting robot positions and games for a session, the OT balanced their clinical judgment of therapeutic appropriateness with the participant’s preferences and current fatigue level, as deemed by the clinician on site. Several design implications follow from the observed selection patterns. First, rehabilitation robot systems should employ a perceptual-accessibility screen administered at intake and physical-demand characterization should also record the type of control a game requires, not only the magnitude or precision of the required movements. TheraStack, for example, is rated at the lowest physical-demand level in our library but requires two distinct, timed button presses coordinated with continuous movement of the same limb. A game can therefore impose low movement demands while still requiring substantial coordination, timing, or control complexity. Representing these interaction requirements separately would distinguish how much movement a game demands from how that movement must be organized to control it.

Second, because games with the same characterized cognitive or physical demand were not selected interchangeably, demand characterization appears more useful for narrowing a library to a clinically appropriate set than for prescribing a single activity. Game libraries should therefore include multiple games within each demand combination so that participant preference can operate within a set of games already judged appropriate by the clinician. A demand combination represented by only one game is fragile. In our library, the only game at the lowest cognitive level also encoded item value by color, so a participant unable to distinguish those colors would have had no low-cognitive-demand alternative. Third, initial preference should be treated as provisional rather than as a basis for permanent exclusion. Additional exposure to the game may change a participants’ enjoyment of it. So, re-offering a rejected activity with supported practice may therefore be more appropriate than removing it immediately from the participant’s rotation.

Fourth, selection of the robot training position should represent constraints that arise from the interaction between the participant and the device, not only from impairment-related demands. One participant was directed away from a robot configuration because their strength in that position could displace the robot base, a safety constraint that the cognitive and physical demand levels do not capture.

Finally, the rationale for each selection should be recorded at the time it is made. A brief structured annotation, such as therapeutic goal, participant preference, fatigue, variety, or safety, would turn the therapist’s judgment from a retrospective explanation into data that could be used to prospectively evaluate demand characterization.

### Dynamic Difficulty Adjustment (DDA) Across the Robotic Gym

A central design requirement of the rehabilitation gym is that people of heterogeneous severity train simultaneously at adjacent stations without a uniform difficulty setting under- or over-challenging any single individual. The clinical data provide converging evidence that the DDA achieved this balance across a wide range of clinical profiles (Table 2).

Mean perceived exertion was light-to-moderate across sessions. Per-game exertion clustered within a narrow range for six of eight games, while Sky Burger and Therastack elicited higher effort consistent with their greater cognitive demand. No game produced exertion near the ceiling, confirming that the DDA maintained a productive effort range across various motor and cognitive demands. NASA-TLX frustration declined to near-floor by the third timepoint, and perceived effort trended downward, consistent with the DDA calibrating to motor improvements over time.

Game-level progression and force-feedback trajectories further illustrate individualized, game-specific adaptation. As reported above, the controller advanced Space Shooter through several difficulty levels for participant 006 while holding Citadel constant, indicating that difficulty was estimated separately for each game rather than from a single participant profile applied across the library. Within the same day, the two games also showed opposite force-feedback trajectories. Citadel’s resistance increased rapidly to near its ceiling and remained there, consistent with mastery of a continuous steering task, whereas Space Shooter’s resistance declined across the session, consistent with accumulating fatigue during a rapid targeting task. Because these opposing trajectories occurred for the same participant on the same day, they suggest that the DDA responded to the interaction between participant’s state and game demand rather than to participant’s state alone.

These findings address the concern raised by Darzi et al. ^23^ that biometric DDA complexity may not yield meaningful benefits over performance-only approaches; the low-frustration profile and progressive level advancement are consistent with the DDA actively moderating challenge in a way a static setting could not replicate across people of differing abilities sharing the same gym floor.

### Social Game Play for Motivation

Although the games are challenging and engaging on their own, prolonged game play could lead to decreased excitement and motivation, such as the novelty effect that we observed. To increase motivation, the therapists encouraged the participants to beat their own high score or another user’s high score. Specifically, participant 028 stayed late for one session to beat the score of participant 029. Similarly, a few users requested that future iterations of the games include a leader board so that they can see their own progress over time and challenge themselves to do better. These requests suggest interest in longitudinal and social forms of feedback beyond the immediate feedback provided during the game. Furthermore, therapists used competition to encourage participants to play more cognitively demanding games (such as SkyBurger). For example, participant 029 had a severe motor deficit, but mild cognitive deficit, and could handle more cognitively loaded games. Although initially reluctant to play the more cognitively demanding game, the therapists encouraged 029 by commenting that many other study participants were typically unable to complete those games. This form of artificial competition appeared to be highly motivating, as they ended up playing that game more than any other game (Figure 5A). These anecdotes support that competition, both with oneself and with others, can serve as motivation to improve game play.

### Clinical Implications

In this study, we demonstrated feasible deployment of a three robot system in a community-based site. Participants are routinely transported to and from the facility, where their day is planned around activities, food, and medical care. Most participants in the center are beyond the acute phase of recovery and do not have assigned therapy. The robotic gym was easily implemented as it became like another “activity” that they could choose to participate in. While this study is limited to deployment in a single community-based site, we note that many community-base sites operate in a similar way. Future versions of the Rehab CARES system could be deployed in other PACE facilities, nursing homes, or senior centers.

As mentioned in the related work, CBR systems have shown success with multiple robotic stations, specifically by allowing treating therapists to see more patients ^20^. Enabling therapists to oversee multiple patients at once is one of the primary benefits of robot-assisted therapy noted in the literature ^28,30^. During this clinical deployment, one OT oversaw one to two participants on the robotic systems and one participant in a separate traditional OT setting. Each participant received 60 minutes of overlapping care, with the traditional OT participant requiring the most one–on–one care. Once participants were set up on the robotic systems, they completed their therapy with minimal supervision. The deployment of an independent, affordable robotic-assisted therapeutic intervention may potentially alleviate some burden in the clinic.

Robot gyms have also been deployed in LMICs ^30^. The Rehab CARES system is unique in that it offers the capacity of a robotic-gym with one device, THEAbot. This affordable, accessible, reconfigurable, 1 DoF system has the capacity to offer a range of upper extremity exercises. By using the same hardware, it reduces the burden on the clinicians to train on multiple systems, while preserving the ability to exercise different muscles within the shoulder, arm, and wrist. Lastly, the gaming platform offers a variety of games with differing motor and cognitive demands. Thus, the CARES system allows the OT to offer motor and cognitive therapy, with minimal supervision ^56^.

### Limitations and Future Directions

Our findings should be interpreted conservatively given the sample size. Despite this, we were able to test cases representative of stroke survivors with varying levels of motor and cognitive impairments. Several limitations constrain the game-level findings. Game selection was uncontrolled and exposure was unequal across games, so these values cannot be compared inferentially. The user experience was rigorously described from the participant’s perspective, while the therapist’s experience of the system was represented by a narrative record compiled during clinical use. Because the gym model depends on one clinician operating several stations concurrently, a therapist-facing user experience and workload would be additionally informative. Perceptual accessibility was not assessed at intake, despite emerging as a recurring barrier and a plausible confounder for performance-driven difficulty adjustment. For example, a participant who cannot distinguish a gold coin from a silver one scores poorly, which may be read as poor motor or cognitive performance rather than perceptual difficulty. In the future, these issues can be mitigated by including adjustable settings for visual components, e.g. color contrast and item size, and redundant options for feedback, e.g. auditory, visual, and cues.

## Conclusion

An affordable, adaptive robotic system for stroke rehabilitation was clinically deployed in a community-based setting. The system was designed to be personalized and to adapt to individual user’s needs across 4 to 6 weeks of therapy. Personalization was achieved through reconfigurable hardware, enabling 3 different movement patterns, and a library of 8 games with distinct motor and cognitive profiles, allowing participants with different motor and cognitive impairments to access therapy. The system was also adapted to support users over time through a DDA system that tailored the challenge of the game and the physical resistance to contemporaneous performance. Across a small, heterogeneous cohort of post-stroke participants, we observed highly individualized utilization patterns with factors such as cognitive and motor demand, personal preferences, therapy goals, and perceptual ability influencing game and position selection. Together, these results suggest successful personalization of robot-assisted therapy using a low-cost, system of 1-DOF robots. We also demonstrated increased usability, decreased frustration, and increased perceived control over time, suggesting that participants grew more comfortable with the system with sustained use. Findings support the clinical viability of an affordable system of robots for personalized, robot-assisted motor/cognitive therapy in real-world community-based settings.

## Acknowledgements

We want to thank Natasha Dastur, OTR/L and Kelly Conway, RN, OTR/L for their clinical expertise in administering robotic rehabilitation and Joana Opong-Duah, Carol Wamsley, DPT and Greg Holt, OTR for conducting patient assessments. We also thank Samuel Gaardsmoe and Saibaba Nargund at Recupero Robotics, LLC for designing and supporting the robot systems and enAblegames™ for designing the therapy games and software architecture. Finally, we thank Mercy LIFE West Philadelphia for providing clinical space and assistance with recruitment and the participants for their enthusiasm.

## Statements and Declarations

### Author contributions

**RMA** coordinated study logistics, collected data, and co-wrote the paper. **YS** developed the DDA system and games, analyzed DDA data, created figures, and co-wrote the paper. **ELW** developed the robotic controller, analyzed user experience data, created figures, and co-wrote the paper. **RJM** characterized the cognitive-motor load of the games and provided clinical oversight. **AJ** designed and directed robot development and provided funding. **PD** led DDA system and game development. **PZC** led recruitment, consented participants, and provided clinical and research oversight. **MJJ** originated project direction and provided funding and resources. All authors reviewed the manuscript.

### Ethical Considerations

This protocol was approved by the Institutional Review Board of the University of Pennsylvania (UPenn) (Protocol no. 853607) and Trinity Health IRB relied on the IRB at UPenn.

### Consent to Participate

All participants gave written informed consent prior to participation in this study (Institutional Review Board number: 853607).

### Consent for Publication

All participants gave written informed consent to publish their results.

### Data Availability

Data is available upon request. Please contact the corresponding author.

